# Diverging trajectories of cardiorenal protection: from early benefit to late attenuation with multifactorial target achievement across cardiovascular-kidney-metabolic syndrome stages

**DOI:** 10.64898/2026.01.14.26344159

**Authors:** Lu Cai, Baiyi Liang, Shiyu Zhou, Huaying Xiao, Ying hu, Hong Ma

## Abstract

**Background:** The extent to which achieving multiple metabolic treatment targets confers sustained cardiorenal protection across all stages of cardiovascular - kidney - metabolic (CKM) syndrome remains uncertain.

**Methods:** This multicenter retrospective cohort study used data from the China Renal Data System (CRDS). Participants were classified into CKM stages 1–4. Metabolic target achievement was defined as simultaneous control of blood pressure (BP<130/80 mmHg), fasting blood glucose (FBG<5.6 mmol/L), and low-density lipoprotein cholesterol (LDL-C<1.8 mmol/L). A metabolic score (0-3) reflected the number of targets met. Outcomes included a composite cardiovascular-renal endpoint and all-cause mortality. Cox and restricted cubic spline regression models were used to assess associations between metabolic control and outcomes in CKM patients.

**Results:** The proportion of patients meeting all three metabolic targets decreased markedly with advancing CKM stage. Kaplan-Meier curves showed increasing risks of cardiovascular-renal events and mortality across higher stages (P<0.001). After adjustment, higher metabolic scores were linked to lower risks: for cardiovascular-renal outcomes, HR 0.82 (95% CI 0.77–0.87) for score 1, 0.72 (0.68–0.77) for score 2, and 0.65 (0.57–0.73) for score 3; for mortality, HR 0.73 (0.62–0.87) for score 1 and 0.68 (0.47–1.00) for score 2 (both P<0.05). However, achieving all three targets did not reduce mortality risk (HR 1.04, 95% CI 0.64–1.68). Stratified analysis showed significant risk reduction in early CKM stages but not in advanced stages. Restricted cubic spline models indicated nonlinear associations between LDL-C and both outcomes, and between FBG, systolic BP, and mortality, after full adjustment (all P<0.05).

**Conclusion:** While multifactorial metabolic target attainment shows strong protective associations with cardiovascular-renal outcomes in early CKM stages, this relationship diminishes in advanced disease. These findings highlight the need for stage-specific management strategies in CKM syndrome.

## Introduction

Cardiovascular diseases (CVD), chronic kidney diseases (CKD) and type 2 diabetes (T2D) are the main causes of death and disability worldwide [1]. For a long time, these diseases have been regarded as separate entities, but more and more evidence indicates that they are interrelated and mutually exacerbate each other under common pathophysiological mechanisms such as obesity, insulin resistance and chronic inflammation [2]. Therefore, the American Heart Association (AHA) officially proposed the concept of Cardiovascular-Kidney-Metabolism (CKM) syndrome in 2023 and released a staging system from stage 0 (without risk factors) to stage 4 (clinical event stage), aiming to integrate management strategies and block this vicious cycle at its source [3].

China is a major burden country for CKM syndrome. Data show that the prevalence of diabetes among Chinese adults has exceeded 11%, while the prevalence of hypertension and chronic kidney diseases is 23.2% and 8.2% respectively [4, 5]. What is more worrying is that the control status of these diseases is not optimistic. Contemporary clinical guidelines emphasize the importance of achieving strict treatment goals to minimize diabetes-related complications. These include maintaining fasting blood glucose (FBG) below 5.6 mmol/L [6, 7], blood pressure (BP) under 130/80 mmHg for individuals with elevated cardiovascular risk or conditions like CKD [7–9], and low-density lipoprotein cholesterol (LDL-C) below 1.8 mmol/L in patients at very high risk for atherosclerotic cardiovascular disease [7,10]. Despite these recommendations, nationwide data reveal that merely 12.4% of diabetic patients meet all three control targets concurrently [11, 12]. This management dilemma is the result of insufficient health literacy of patients, imperfect hierarchical diagnosis and treatment in the medical system, and social and cultural factors.

Although the CKM staging system offers a theoretical framework for risk stratification, evidence on how Chinese patients achieve treatment targets across different stages in real-world clinical practice remains limited. The extent of prognostic differences among patients at various CKM stages and whether the protective effect of achieving metabolic goals varies by stage is still unclear due to a lack of large-scale studies. Therefore, this study aims to systematically characterize the treatment target attainment (FBG <5.6 mmol/L, BP <130/80 mmHg, and LDL-C <1.8 mmol/L) landscape in China’s CKM population using a multicenter cohort and to quantitatively assess the association between achievement of key metabolic indicators and cardiovascular-kidney outcomes, thereby providing robust data to inform precision-based, stage-specific CKM management strategies.

## Methods

### Statement of Ethics

The study protocol was approved by the Medical Ethics Committee of Nanfang Hospital, Southern Medical University (approval number: NFEC-2019-213), which waived the requirement for patient informed consent due to the retrospective nature of the study. This study was also approved by the China Office of Human Genetic Resources for Data Preservation Application (approval number: 2021-BC0037) and was performed in accordance with the Strengthening the Reporting of Observational Studies in Epidemiology (STROBE) guidelines [13].

### Study Design, Population, and Data Source

This study is a large, multicenter, retrospective cohort study. Patients with metabolic syndrome aged over 18 years old were enrolled from January 4,2000 to December 12, 2024 in the China Renal Data System (CRDS). For every eligible participant, the index date was established as the date of their initial clinical visit during the study period that fulfilled all the inclusion criteria. The baseline period was defined as the 12-month interval immediately preceding and including the index date. Using the China Renal Data System which has been described in our previous study [14-16], we conducted a large multicenter retrospective study cohort of 7084339 patients hospitalized at 32 medical centers throughout China from 1 January 2000 to 26 December 2024. In briefly, this database contains information on these patients, both while in hospital and as outpatients. Data recorded included patients’demographic characteristics, such as age, sex, smoking and drinking status; clinical characteristics, such as blood pressure, vital signs, diagnosis codes at admission and discharge, dates of diagnosis and drugs prescribed; surgical information, such as dates of operation and operation procedure codes; and laboratory test results, including the time of testing.Prescription data included names, codes, doses, dose units, frequency, route, and start and stop times of all drugs. The data from all participating hospitals were pooled and analyzed at the National Clinical Research Center for Kidney Disease in Guangzhou, China. All the laboratories of the participating hospitals had passed the annual External Quality Assessment of the Chinese National Center for Clinical Laboratories.

### Inclusion criteria

(1) Age≥18 years;

(2) Presence of at least three of the following five components metabolic abnormality as defined by the International Diabetes Federation (IDF) criteria for metabolic syndrome. Metabolic syndrome was defined according to the IDF standard [17], requiring fulfillment of at least three of the following five components: abdominal obesity (waist circumference ≥90 cm in men or ≥80 cm in women); hypertension [systolic blood pressure (SBP) ≥130 mmHg and/or diastolic blood pressure (DBP) ≥85 mmHg, or current use of antihypertensive medication);elevated triglycerides (TG) (TG ≥1.7 mmol/L or on specific treatment); reduced high-density lipoprotein cholesterol (HDL-C) (HDL-C <1.0 mmol/L in men and <1.3 mmol/L in women, or on lipid-lowering therapy); and FBG (FBG≥5.6 mmol/L or previously diagnosed with type 2 diabetes).

### Exclusion criteria

(1) Malignancy with life expectancy <1 year; (2) Secondary hypertension or diabetes defined as Cushing’s syndrome, pheochromocytoma and based on the ICD-10 coding system including E22.0, E22.001, E26.0, E26.1, E26.8, E26.9, E03.0-E03.05, E03.8, E03.9, E05.0-E05.5, E05.8 and E05.9; (3) decompensated liver cirrhosis; (4) patients without data of BMI, waist circumference, BP, TG, LDL-C, HLD-C and FBG; (5) patients whose outcomes occurred before the baseline.

Finally 78890 patients were enrolled in this study (Figure 1).

**Figure 1.**
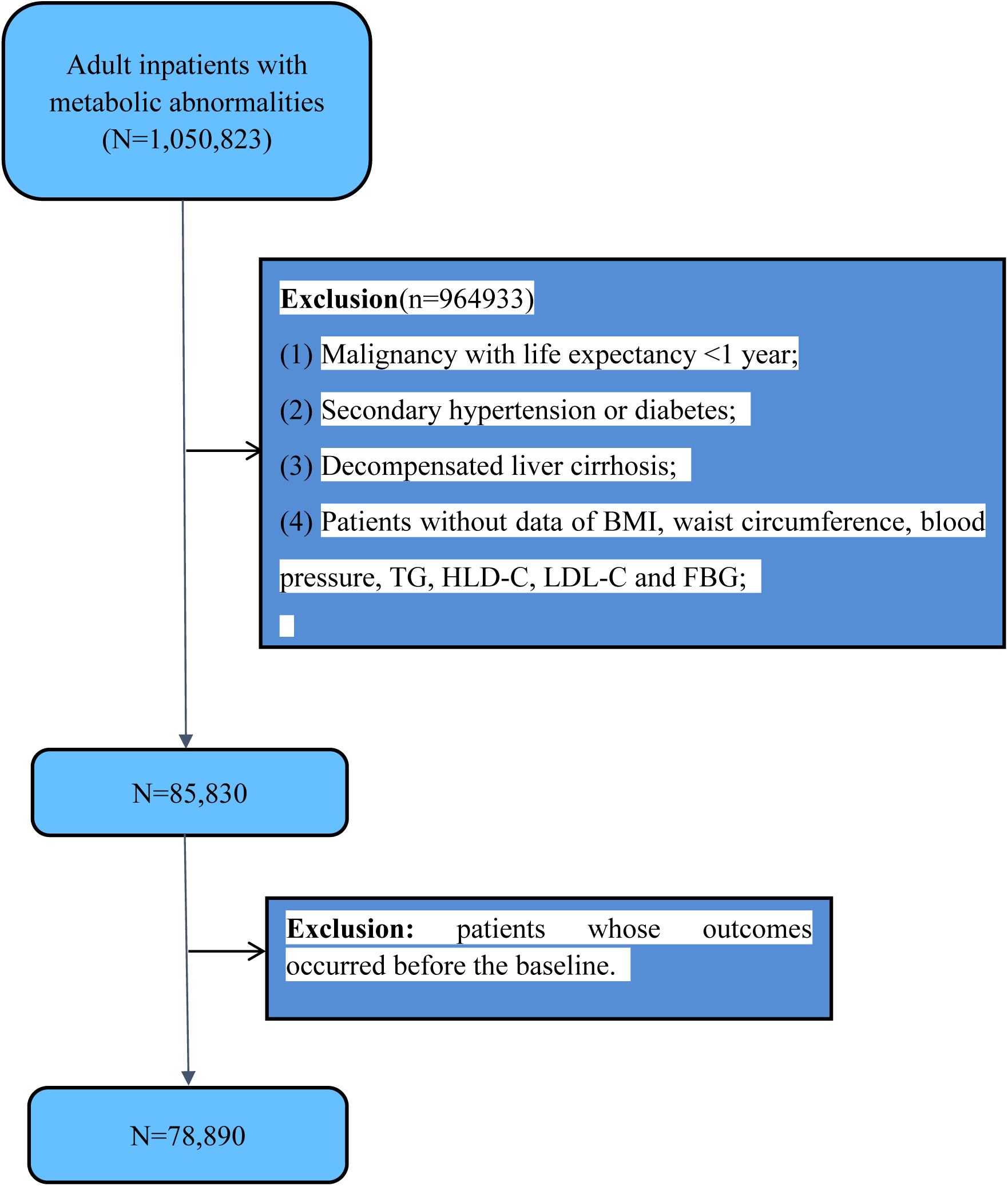
Flow chart for this retrospective study. Abbreviations: CVD, cardiovascular diseases; BMI, body mass index; TG: total triglycerides; HDL-C: high-density lipoprotein cholesterol; LDL-C: low-density lipoprotein cholesterol; FBG, fasting blood glucose.

### Deffnition of CKM syndrome stages 1 to 4

Based on the AHA Presidential Advisory Statement [2], the stages of CKM syndrome are categorized as follows: Stage 0 refers to the absence of any CKM risk factors. Stage 1 is characterized by abnormal or impaired adipose tissue function. Stage 2 involves the presence of metabolic abnormalities—such as type 2 diabetes, hypertension, elevated triglyceride levels—or chronic kidney disease. Stage 3 indicates subclinical cardiovascular disease occurring within the framework of CKM syndrome. Stage 4 represents clinically evident cardiovascular disease, encompassing conditions such as coronary artery disease, heart failure, stroke, peripheral arterial disease, and atrial fibrillation, all in the context of CKM syndrome.

### Metabolic score

We defined a metabolic score (ranging from 0 to 3) as the sum of the following treatment targets achieved at the baseline: (1) FBG <5.6 mmol/L; (2) BP <130/80 mmHg; and (3) LDL-C <1.8 mmol/L. A score of 0 indicates no targets met, 1 indicates one target met, 2 indicates two targets met, and 3 indicates all three targets met.

### Definition of Baseline and Index Date

For each eligible participant, the index date was the first clinical visit during the study period that met all inclusion criteria. The baseline period covered the 12 months before and including this date. Baseline characteristics were defined as follows:

1. Pre-existing comorbidities were identified from healthcare records during the 12-month baseline period.
2. Laboratory values and vital signs were taken from the latest available measurement within baseline. If unavailable, the first measurement within one month after the index date was used.
3. All baseline data were collected before any outcome event to ensure temporal validity.

### Data collection and laboratory measurements

A medical history and findings on physical examination were recorded. Hypertension, diabetes (DM) and Chronic obstructive pulmonary disease (COPD) were defined as a clinical history of the diagnoses recorded in patients’ notes.

Baseline biochemical parameters were collected at the baseline, including blood pressure (BP), height, weight, body mass index (BMI) hemoglobin (Hb), hypersensitive c-reactive proteinhs-CRP (hs-CRP), albumin (ALB), urinary protein (upro), TG, total cholesterol (TCHO), LDL-C, HDL-C, uric acid (ua), urea nitrogen (BUN), serum creatinine (Cr), alanine aminotransferase (ALT), aspartate aminotransferase (AST), direct bilirubin (DBIL), indirect bilirubin (IBIL), Calcium (Ca), potassium (K), Hemoglobin A1c (HbA1c) and fasting blood glucose (FBG). Glomerular filtration rate was estimated using the Chronic Kidney Disease Epidemiology Collaboration (CKD-EPI) formula [18].

### The endpoint

The primary end point is cardiovascular-renal composite outcome (first occurrence of any of the following events). Cardiovascular events refer to myocardial infarction, defined according to the ICD-10 codes I21.0–I21.4, I21.9, I22.0, I22.1, I22.8, I22.9, I23.0–I23.3, and I23.6, as well as ischemic stroke, defined by ICD-10 codes I63.0– I63.9. Renal event was defined as eGFR reduction of 40% or progression to end-stage renal disease (ESRD). ESRD was identified as eGFR<15ml/min/1.73m^2^, maintenance dialysis or renal transplantation. The second endpoint is all-cause mortality. The follow-up period began at the index date and extended to the earliest of the following: the first occurrence of the endpoint, the date of the last documented clinical encounter in the database, or the end of the study observation period.

### Statistical analysis

The data were presented as mean ± standard deviation for normally distributed continuous variables, and number (proportion) for categorical variables. Student’t test was used for comparison between groups; The measurement data of non normal distribution were expressed by median (interquartile interval), and the comparison between groups was performed by Wilcox rank sum test.

Cox proportional hazards regression was performed to assess the association between achievement of metabolic targets and the risk of a composite cardiovascular-renal endpoint as well as all-cause mortality. The Kaplan-Meier method was used to estimate the cumulative incidence of the composite outcome and all-cause mortality across different CKM stages, with group differences compared using the log-rank test. Restricted cubic spline (RCS) analysis was applied to model the potential non-linear associations between continuous metabolic variables and both the composite cardiovascular-renal outcome and all-cause mortality, enabling a more flexible evaluation of dose-response relationships.

The Cox proportional hazards model was employed to assess whether the association between metabolic indicators and outcomes varied across CKM stages within subgroups stratified by COPD comorbidity, and baseline eGFR category (≥60 vs. <60 ml/min/1.73 m²). Forest plots were generated to visualize the effect estimates and corresponding 95% confidence intervals for each subgroup, facilitating a clear and intuitive comparison of heterogeneity across strata. To account for the competing risk of death, the Fine-Gray subdistribution hazard model was applied; multiple imputation using the MICE package was performed to address missing data.

A 2-tailed p<0.05 was considered statistically significant in all analyses. All statistical analyses were performed using R software, version (4.3.1).

## Results

### Baseline demographic and clinical characteristics of the study population

Of the 78890 patients with an average age of (51.63 ± 15.44) years at recruitment were enrolled. Among them, 39754 (50.39%) were female. The participants met criteria for CKM syndrome, with 36.97%, 62.42%, 0.44%, and 0.17% classified as CKM stages 1, 2, 3, and 4, respectively. As the CKM syndrome progresses through its stages, the proportion of patients achieving simultaneous control of BP, FBG and LDL-C drops sharply. Baseline characteristics of patients with CKM are shown in Table 1.

**Table 1.** Baseline demographic and clinical characteristics of the study population.

| CKM Stage |  | 1 | 2 | 3 | 4 | p |
| --- | --- | --- | --- | --- | --- | --- |
| n |  | 29163 | 49245 | 349 | 133 |  |
| Sex (%) | Female | 18389 ( 63.06) | 21154 ( 42.96) | 149 ( 42.69) | 62 ( 46.62) | <0.001 |
|  | Male | 10774 ( 36.94) | 28091 ( 57.04) | 200 ( 57.31) | 71 ( 53.38) |  |
| Age, years |  | 47.32 (16.65) | 54.07 (14.06) | 59.50 (13.63) | 68.30 (14.21) | <0.001 |
| Heart rate, beats/minute |  | 80.88 (11.01) | 81.40 (11.50) | 84.08 (13.23) | 79.23 (11.08) | <0.001 |
| SBP, mmHg |  | 123.83 (17.95) | 129.37 (18.55) | 145.40 (22.77) | 144.44 (25.30) | <0.001 |
| DBP, mmHg |  | 75.74 (11.33) | 79.23 (37.95) | 81.90 (13.08) | 76.12 (16.65) | <0.001 |
| Height, m |  | 1.64 (0.07) | 1.66 (0.08) | 1.64 (0.09) | 1.63 (0.09) | <0.001 |
| Weight, kg |  | 68.69 (13.30) | 72.78 (14.58) | 67.73 (14.14) | 66.32 (15.25) | <0.001 |
| BMI, kg/m<sup>2</sup> |  | 25.21 (4.10) | 26.07 (4.13) | 24.92 (4.14) | 25.01 (4.93) | <0.001 |
| COPD (%) |  | 573 (1.97) | 1787 (3.63) | 18 (5.16) | 25 ( 18.80) | <0.001 |
| hypertension (%) |  | 0 (0.00) | 11512 (23.38) | 177 (50.72) | 117 (87.97) | <0.001 |
| diabetes(%) |  | 0 (0.00) | 25500 (51.78) | 200 (57.31) | 91 (68.42) | <0.001 |
| hs-CRP, mg/L |  | 27.92 (1126.64) | 10.30 (28.62) | 17.48 (38.23) | 23.62 (40.59) | 0.332 |
| Hb, g/L |  | 130.16 (19.85) | 139.76 (20.55) | 102.99 (23.25) | 100.71 (19.06) | <0.001 |
| TCHO,mmol/L |  | 4.98 (1.46) | 4.66 (1.36) | 4.84 (1.63) | 4.40 (1.43) | <0.001 |
| TG,mmol/L |  | 1.94 (1.29) | 2.47 (2.39) | 2.24 (1.83) | 2.10 (2.03) | <0.001 |
| LDLC,mmol/L |  | 2.97 (1.00) | 2.76 (0.95) | 2.84 (1.22) | 2.45 (1.01) | <0.001 |
| HDLc,mmol/L |  | 1.41 (0.48) | 1.07 (0.34) | 1.06 (0.38) | 1.02 (0.36) | <0.001 |
| UA, umol/L |  | 312.83 (103.15) | 343.95 (134.06) | 454.42 (132.13) | 461.32 (131.76) | <0.001 |
| Cr, umol/L |  | 61.26 (35.23) | 68.75 (34.03) | 352.68 (240.61) | 280.62 (136.42) | <0.001 |
| eGFR, ml/min/1.73 m<sup>2</sup> |  | 107.885 (22.99) | 98.483 (21.97) | 18.375 (7.76) | 19.897 (6.75) | <0.001 |
| Ca, mmol/L |  | 2.25 (0.14) | 2.27 (0.14) | 2.15 (0.19) | 2.14 (0.18) | <0.001 |
| P, mmol/L |  | 1.17 (0.22) | 1.17 (0.23) | 1.43 (0.43) | 1.36 (0.30) | <0.001 |
| K, mmo/L |  | 3.88 (0.39) | 3.92 (0.44) | 4.40 (0.76) | 4.34 (0.63) | <0.001 |
| upro (%) | - | 20982 (81.16) | 34089 (77.63) | 31 (10.23) | 24 (20.00) | <0.001 |
|  | ± | 3058 (11.83) | 4220 (9.61) | 16 (5.28) | 14 (11.67) |  |
|  | 1+ | 1230 (4.76) | 3035 (6.91) | 44 (14.52) | 17 (14.17) |  |
|  | 2+ | 381 (1.47) | 1617 (3.68) | 98 (32.34) | 26 (21.67) |  |
|  | 3+ | 193 (0.75) | 883 (2.01) | 104 (34.32) | 34 (28.33) |  |
|  | 4+ | 8 (0.03) | 70 (0.16) | 10 (3.30) | 5 (4.17) |  |
| UPCR ,mg/g |  | 1178.99 (8371.28) | 930.86 (13184.54) | 3545.96 (3681.75) | 3954.95 (3564.02) | 0.397 |
| ALT, U/L |  | 25.07 (56.45) | 30.12 (58.14) | 20.15 (22.06) | 18.27 (13.50) | <0.001 |
| AST, U/L |  | 25.70 (70.36) | 26.66 (75.80) | 22.32 (20.25) | 23.99 (20.50) | 0.258 |
| DBIL, umol/L |  | 2.73 (5.26) | 3.60 (7.12) | 2.21 (1.83) | 2.86 (5.36) | <0.001 |
| IBIL, umol/L |  | 8.84 (5.52) | 9.24 (6.04) | 5.35 (3.41) | 5.86 (6.30) | <0.001 |
| FBG, mmol/L |  | 5.60 (2.44) | 7.79 (3.52) | 7.18 (4.34) | 6.58 (2.69) | <0.001 |
| HbA1c, % |  | 6.94 (2.48) | 8.71 (2.64) | 7.60 (2.13) | 7.39 (1.95) | <0.001 |
| Metabolic score (%) | 0 | 1770 ( 6.07) | 10267 (20.85) | 80 (22.92) | 10 ( 7.52) | <0.001 |
|  | 1 | 8046 (27.59) | 24936 (50.64) | 151 (43.27) | 64 (48.12) |  |
|  | 2 | 17709 (60.72) | 12643 (25.67) | 101 (28.94) | 46 (34.59) |  |
|  | 3 | 1638 ( 5.62) | 1399 ( 2.84) | 17 ( 4.87) | 13 ( 9.77) |  |
| Total stay,days |  | 3.85 (16.14) | 4.88 (9.27) | 7.75 (14.78) | 6.95 (13.81) | <0.001 |
| Time to outcome,years |  | 0.78 (1.43) | 1.05 (1.62) | 0.83 (1.34) | 1.07 (1.43) | <0.001 |
| death (%) |  | 294 (1.01) | 953 (1.94) | 42 (12.03) | 22 (16.54) | <0.001 |

Abbreviations: SBP: systolic blood pressure; DBP: diastolic blood pressure;TG: total triglycerides; UA: uric acid; TCHO: total cholesterol; HDL-C: high-density lipoprotein cholesterol; LDL-C: low-density lipoprotein cholesterol; Cr: creatinine; ALT: alanine aminotransferase; AST: aspartate aminotransferase; DBIL: direct bilirubin; IBIL: indirect bilirubin; Ca: calcium; K: potassium; P:Phosphorus; FBG:fasting blood glucose; HbA1c: Hemoglobin A1c; COPD: Chronic obstructive pulmonary disease;Total stay: hospital stay from admission to discharge;Time to outcome: Time from admission to the occurrence of the outcome for patients with major/minor outcomes or the time of the last follow-up record for patients without any outcome; Metabolic score 0 indicates no targets met, 1 indicates one target met, 2 indicates two targets met, and 3 indicates all three targets met.

### Metabolic score and prognosis

The Kaplan-Meier curve showed as CKM stage advances, the risk of experiencing the cardiovascular-renal composite outcome (log-rank test, P<0.001, Figure. 2A) and all-cause mortality (log-rank test, P<0.001, Figure. 2B) progressively increases among patients.

**Figure 2.**
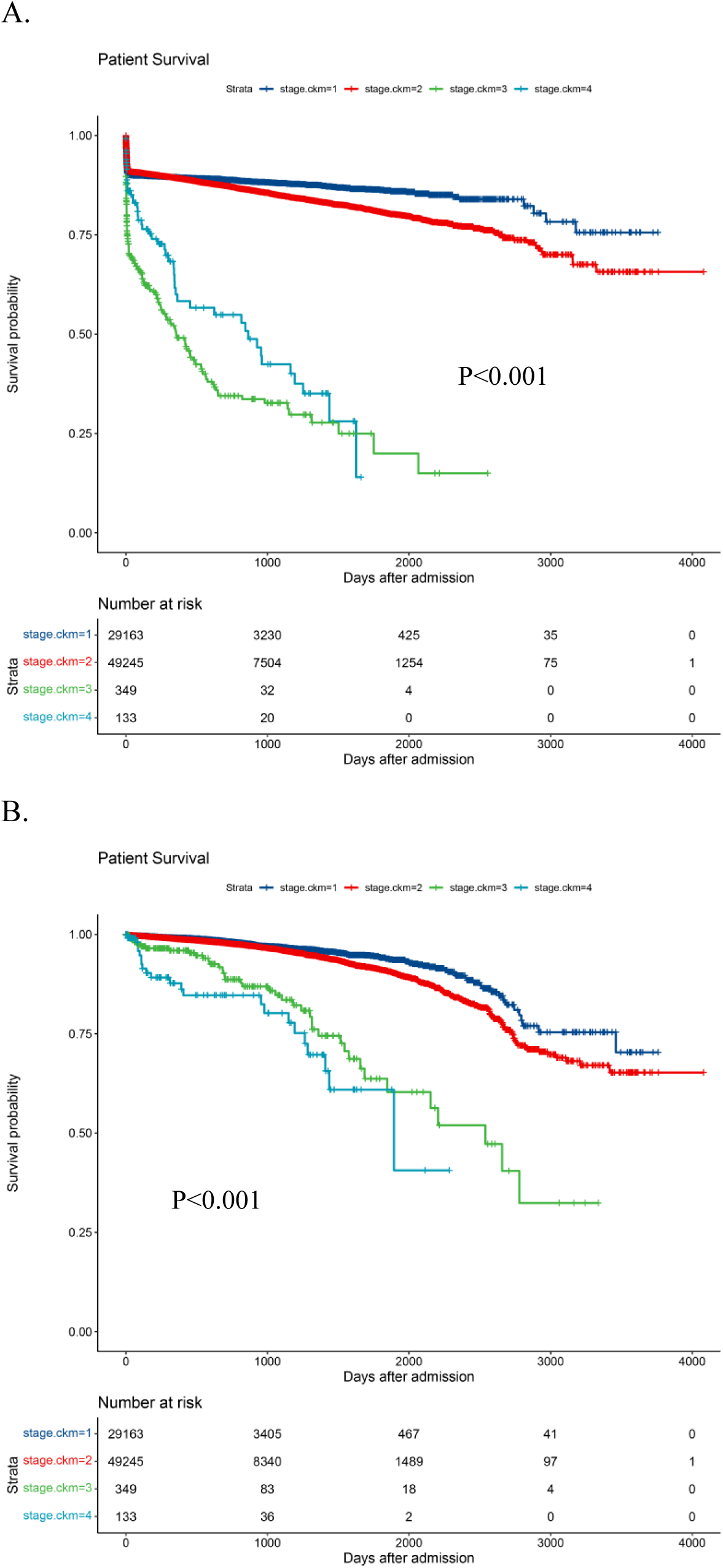
Kaplan-Meier estimates of (A) cardiovascular-renal survival rate and (B) all-cause survival rate according to CKM stage.

After adjusting for covariates mentioned above and clinical significance including age, sex, BMI, CKM stage, eGFR, TCHO, TG, HDLC, upro, COPD, diabetes and hypertension, as the number of achieved metabolic targets increases, improved metabolic status is increasingly associated with a lower risk of cardiovascular-renal composite endpoints [metabolic score 1: hazard ratio (HR) 0.82, 95% confidence interval (CI) 0.77-0.87, P < 0.05; metabolic score 2: HR 0.72, 95% CI 0.68-0.77, P < 0.05; metabolic score 3:HR 0.65, 95% CI 0.57-0.73, P < 0.05] and all-cause mortality (metabolic score 1: HR 0.73, 95%CI 0.62-0.87, P < 0.05; metabolic score 2: HR 0.68, 95% CI 0.47-1.00, P < 0.05) (Table 3). However, this protective relationship was attenuated and did not reach statistical significance for patients achieving metabolic score 3 (HR 1.04, 95% CI 0.64-1.68, P =0.876) (Table 3).

**Table 2:** Multivariate cox proportional analysis of metabolic score to predict cardiovascular-renal composite outcome and all-cause mortality.

| Outcome | Num of Event / N | HR(95%CI) | P |
| --- | --- | --- | --- |
| **Cardiovascular-renal composite outcome** |  |  |  |
| Metabolic score 0 | 1620 / 12127 | 1.00 (Ref.) | - |
| Metabolic score 1 | 3502 / 33197 | 0.82 (0.77-0.87) | <0.05 |
| Metabolic score 2 | 2410 / 30499 | 0.72 (0.68-0.77) | <0.05 |
| Metabolic score 3 | 397 / 3067 | 0.65 (0.57-0.73) | <0.05 |
| **All-cause mortality** |  |  |  |
| Metabolic score 0 | 290 / 12127 | 1.00 (Ref.) | - |
| Metabolic score 1 | 595 / 33197 | 0.73 (0.62-0.87) | <0.05 |
| Metabolic score 2 | 366 / 30499 | 0.68 (0.47-1.00) | <0.05 |
| Metabolic score 3 | 60 / 3067 | 1.04 (0.64-1.68) | 0.876 |

Abbreviations: CKM: cardiovascular – kidney – metabolic syndrome; Num: Number; HR: hazard ratio; CI: confidence interval; Ref: reference. Hazard ratios were adjusted for age, sex, BMI, CKM stage, eGFR, TCHO, TG, HDLC, upro, COPD, diabetes and hypertension. Other Abbreviations as in Table 1.

**Table 3:**
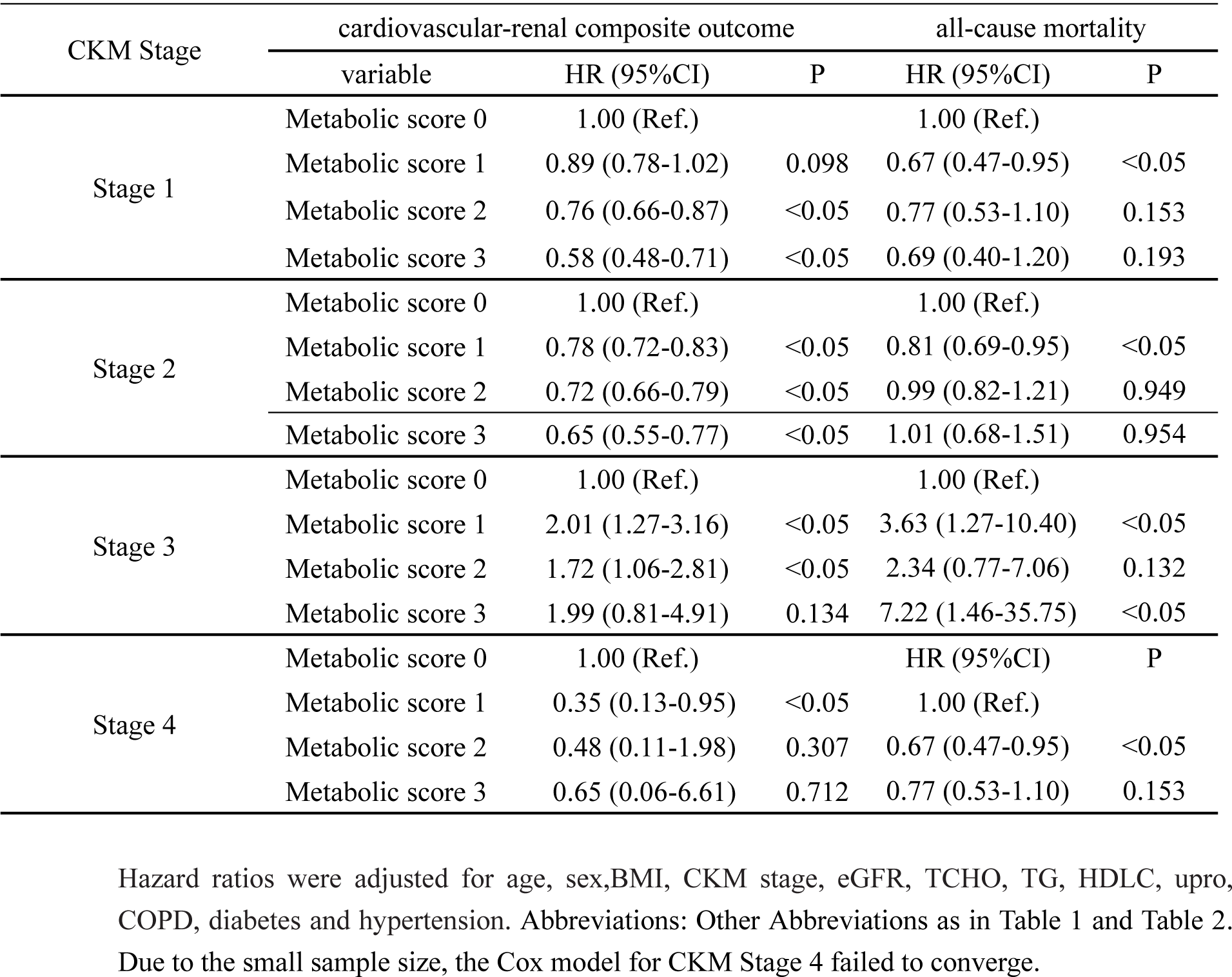
Hazard ratios for risks of cardiovascular-renal composite outcome and all-cause mortality stratified by cardiovascular–kidney–metabolic syndrome stage.

In CKM stages 1 and 2, CKM staging significantly modified the association between metabolic health and clinical outcome risk: as the number of metabolic indicators meeting the threshold increased, improved metabolic status was progressively associated with a lower risk of the cardiovascular-renal composite endpoint (CKM stage 1:metabolic score 1: HR 0.89, 95%CI 0.78-1.02, P=0.098; metabolic score 2: HR 0.76, 95% CI 0.66-0.87, P < 0.05; metabolic score 3:HR 0.58, 95% CI 0.48-0.71, P < 0.05; CKM stage 2:metabolic score 1: HR 0.78, 95%CI 0.72-0.83, P<0.05; metabolic score 2: HR 0.72, 95% CI 0.66-0.79, P < 0.05; metabolic score 3:HR 0.65, 95% CI 0.55-0.77, P < 0.05) (Table 3). In contrast, in CKM stages 3 and 4, the limited sample size prevented the identification of statistically significant associations (Table 3).

A growing number of metabolic indicators surpassing the predefined threshold was not linked to markedly improved metabolic health and a substantially diminished risk of all-cause mortality. Due to the small sample size, the Cox model for CKM Stage 4 failed to converge (Table 3).

### Subgroup analysis

In subgroup analysis, the association between metabolic score and cardiovascular-renal composite outcome as well as all-cause mortality were not different in patients with or without eGFR <60ml/min/1.72m^2^ and COPD (P for interaction > 0.05) (Figure 3A, 3B).

**Figure 3.**
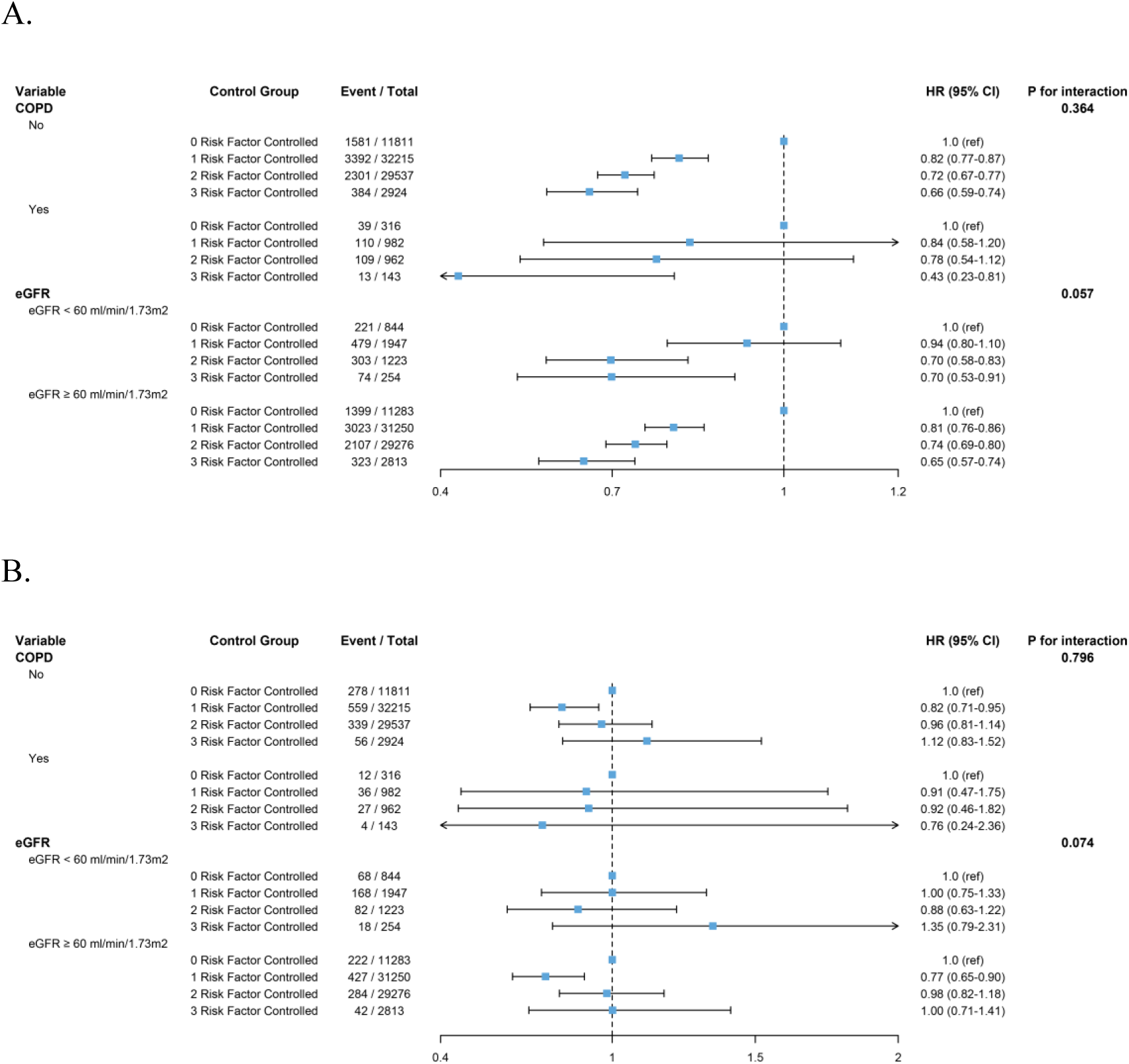
The association between metabolic score and (A) cardiovascular-renal composite outcome as well as (B) all-cause mortality in different subgroups using Cox regression models. Abbreviations as in Table 1, and adjusted variables as in Table 2.

### Non-linear associations of baseline treatment targets with outcomes

Restricted cubic spline (RCS) Cox proportional hazard regression models were used to further examine the association between FBG, LDL-C, SBP and the risk of cardiovascular-renal composite outcome as well as all-cause mortality among patients with CKM syndrome. These RCS curves demonstrated a signiffcantly non-linear relationship between LDL-C and cardiovascular-renal composite outcome in Figure 4B as well as all-cause mortality in Figure 4E, FBG as well as SBP and all-cause mortality in Figure 4D and Figure 4F with full adjustment for covariates (all P<0.05). A linear relationship between LDL-C as well as systolic BP (SBP) and cardiovascular-renal composite outcome in Figure 4A in Figure 4C, was observed both with covariate adjustment (P for non-linear > 0.05).

**Figure 4.**
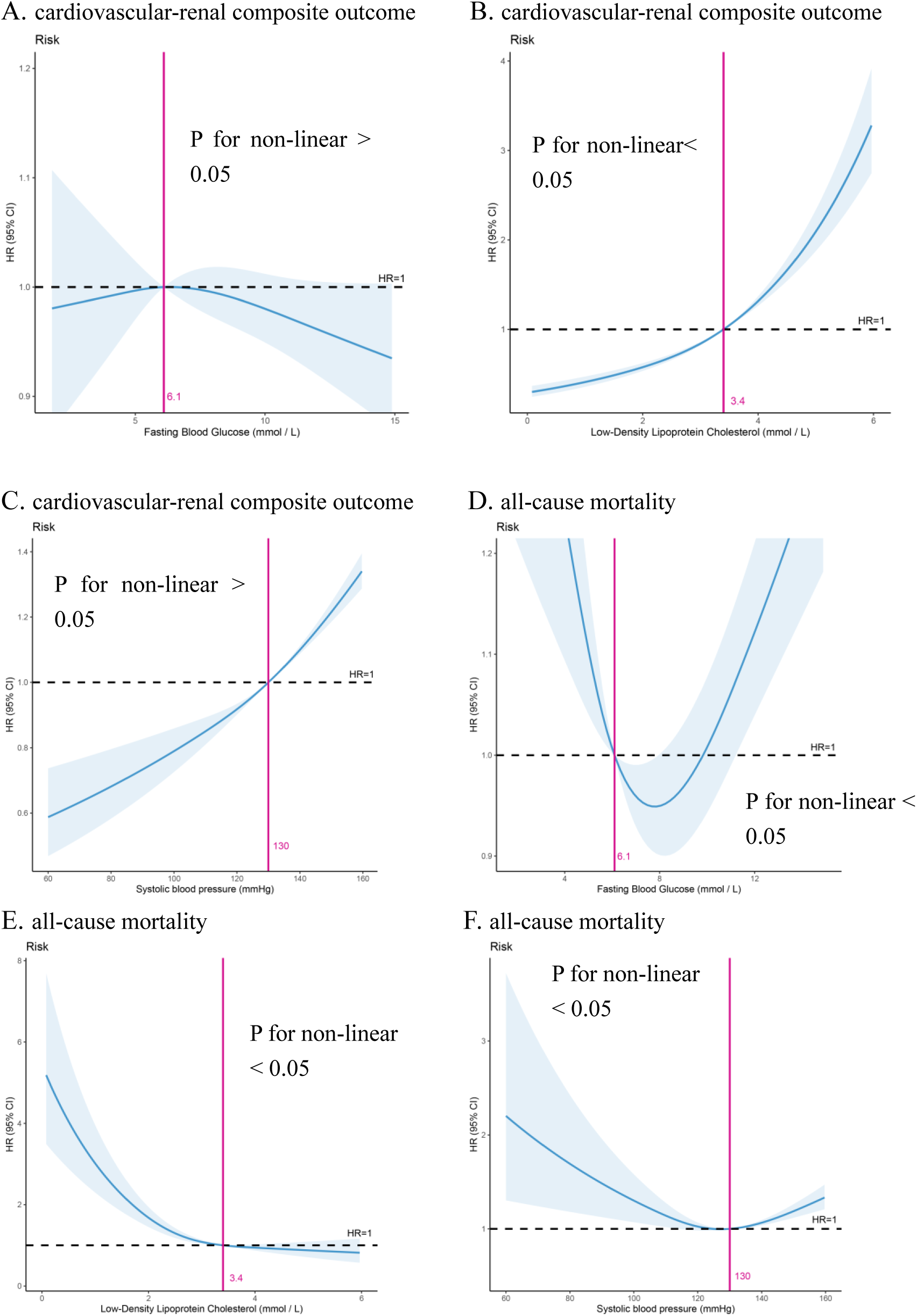
The RCS analysis between (A) FBG and cardiovascular-renal composite outcome, (B) LDL-C and cardiovascular-renal composite outcome, (C) SBP and cardiovascular-renal composite outcome, (D) FBG and all-cause mortality, (E) LDL-C and all-cause mortality, (F) SBP and all-cause mortality, in patients with CKM syndrome. Abbreviations: RCS: Restricted cubic spline. Other abbreviations as in Table 1, and Adjusted variables as in Table 2.

## Discussion

This large, multicenter retrospective cohort study provides a comprehensive analysis of the complex interplay between CKM syndrome staging, multifactorial metabolic target attainment, and clinical outcomes. Our principal findings reveal three critical insights: first, a striking inverse relationship exists between CKM disease progression and the achievement of combined treatment targets; second, a graded protective association between the number of metabolic targets achieved and cardiovascular-renal risk is evident in early CKM stages but attenuates notably in advanced stages, particularly for all-cause mortality; and third, the relationship patterns of traditional risk factors (FBG, LDL-C, SBP) with different outcomes in the CKM spectrum vary, including both linear and non-linear associations.

The steep decline in the proportion of patients achieving simultaneous control of BP, FBG, and LDL-C as CKM stage advances underscores a profound clinical care gap. This aligns with the concept of "therapeutic inertia" magnified by disease complexity [19]. In stages 1 and 2, where pathophysiological processes may be more modifiable, we observed a clear dose-response relationship: each additionally achieved metabolic target was associated with a stepwise reduction in the risk of the cardiovascular-renal composite endpoint. This reinforces the paramount importance of aggressive, multifactorial intervention early in the CKM continuum, as recommended by recent guidelines [20]. The strength of this association, even after extensive adjustment for covariates including baseline eGFR and proteinuria, suggests that the benefits of comprehensive metabolic control are substantial and independent.

However, the attenuation of this protective effect in advanced CKM stages, and its complete loss for all-cause mortality in patients achieving all three targets (Metabolic Score 3), presents a critical nuance. This divergence between the composite endpoint and mortality risk in the most controlled group warrants careful interpretation. Several non-mutually exclusive hypotheses may explain this finding. First, it may reflect the phenomenon of "competing risks" and reverse causality. Patients in stages 3 and 4 often carry a high burden of non-cardiometabolic comorbidities and established, potentially irreversible organ damage. In this context, the progression of pre-existing heart failure, terminal kidney failure, or other non-cardiometabolic conditions may drive mortality, thereby diminishing the measurable protective effect of metabolic control on death [21]. Second, the small sample size in advanced stages, particularly stage 4 where the model failed to converge, inevitably limits statistical power and the precision of our risk estimates[22]. Third, the "floor effect" of risk factors in late-stage disease and the potential for treatment-related adverse effects in a frail population could also contribute to this observed attenuation [23].

Our study uncovers a more complex pattern in how traditional cardiometabolic risk factors relate to various adverse outcomes across the spectrum of chronic kidney disease. Although low-density lipoprotein cholesterol and systolic blood pressure show a linear dose-response association with the composite cardiovascular-renal outcome—aligning with the conventional "lower is better" principle for this particular endpoint [24, 25]—their relationships with all-cause mortality, as well as that of fasting blood glucose, are clearly nonlinear [26–28]. This nonlinearity points to possible threshold effects or J-shaped associations with mortality, implying that the biological pathways connecting extreme levels of these markers to increased death risk may go beyond atherosclerosis and include competing risks such as frailty, undernutrition, and non-cardiovascular causes [29, 30]. As a result, the assumption of a consistent linear association between traditional risk factors and clinical outcomes cannot be generalized across all endpoints in individuals with CKD. These observations highlight the importance of developing outcome-specific risk management approaches and emphasize the intricate balance between cardiovascular and non-cardiovascular mechanisms influencing survival in this population [31].

Our subgroup analysis indicating no effect modification by baseline eGFR or COPD status suggests that the relationship between metabolic target attainment and outcomes is consistent across these important patient subgroups. This further strengthens the generalizability of the core finding that achieving more targets is beneficial in the early-to-mid CKM stages.

Several limitations must be acknowledged. The observational design precludes definitive causal inferences. The retrospective nature of the data may introduce unmeasured confounding, such as medication adherence, diet, and physical activity. The stark imbalance in sample size across CKM stages, with very few patients in stages 3 and 4, is a major limitation for drawing firm conclusions about these advanced stages. Finally, the use of a single baseline measurement for metabolic targets does not account for their variability over time.

In conclusion, our findings delineate a dual clinical imperative. For the vast majority of patients in CKM stages 1 and 2, our data provide powerful real-world evidence supporting intensive, guideline-directed management of all three metabolic domains to mitigate cardiovascular-renal risk. Conversely, for the challenging population with advanced CKM syndrome, a singular focus on these traditional metabolic targets may be insufficient to improve survival. Future management strategies for these patients must evolve to embrace a more holistic, patient-centered approach that integrates metabolic control with the management of advanced organ dysfunction, frailty, and polypharmacy, a paradigm that future prospective studies in well-characterized cohorts must urgently explore.

## Author Contributions

Lu Cai proposed the design of work, analysed, drafted, revised the manuscript and provided final supervision. Boyi Liang and Shiyu Zhou provided efforts in data analysis. Lu Cai, Huaying Xiao,Ying Hu and Hong Ma conceived and designed the study. All authors proofread the manuscript critically and approved the final version of the manuscript to be published.

## Data Availability Statement

The data underlying this article will be shared on reasonable request to the corresponding author.

## References

[1] GBD 2021 Diseases and Injuries Collaborators. Global burden of 369 diseases and injuries in 204 countries and territories, 1990-2019: a systematic analysis for the Global Burden of Disease Study 2019. Lancet. 2020, 17;396(10258):1204-1222.

[2] Ndumele CE, Rangaswami J, Chow SL, et al. Cardiovascular-Kidney-Metabolic Health: A Presidential Advisory From the American Heart Association. Circulation. 2023;148(20):1606–1635.

[3] Ndumele CE, Neeland IJ, Tuttle KR, et al. A Synopsis of the Evidence for the Science and Clinical Management of Cardiovascular-Kidney-Metabolic (CKM) Syndrome: A Scientific Statement From the American Heart Association. Circulation. 2023;148(20):1636–1664.

[4] Li Y, Teng D, Shi X, et al. Prevalence of diabetes recorded in mainland China using 2018 diagnostic criteria from the American Diabetes Association: national cross sectional study. BMJ. 2020; 369: m997.

[5] Zhang L, Wang F, Wang L, et al. Prevalence of chronic kidney disease in China: a cross-sectional survey. Lancet. 2012;379(9818):815–822.

[6] American Diabetes Association Professional Practice Committee. 2. Diagnosis and Classification of Diabetes: Standards of Care in Diabetes-2025. Diabetes Care. 2025; 48(1 Suppl 1):S27-S49.

[7] Kidney Disease: Improving Global Outcomes (KDIGO) Diabetes Work Group. KDIGO 2022 Clinical Practice Guideline for Diabetes Management in Chronic Kidney Disease. Kidney Int. 2022;102(5S):S1-S127.

[8] Whelton PK, Carey RM, Aronow WS, et al. 2017 ACC/ AHA / AAPA /ABC /ACPM/AGS/APhA/ASH/ASPC/NMA/PCNA Guideline forthe Prevention, Detection, Evaluation,and Management of High Blood Pressure in Adults:Executive Summary:A Report ofthe American College of Cardiology/American Heart Association Task Force on Clinical Practice Guidelines. Hypertension.2018;71(6):1269-1324.

[9] Ioannidou E, Shabnam S, Abner S, et al. Effect of more versus less intensive blood pressure control on cardiovascular, renal and mortality outcomes in people with type 2 diabetes: A systematic review and meta-analysis. Diabetes Metab Syndr. 2023;17(6):102782.

[10] Mach F, Baigent C, Catapano AL, et al. 2019 ESC/EAS Guidelines for the management of dyslipidaemias: lipid modification to reduce cardiovascular risk. Eur Heart J. 2020;41(1):111-188.

[11] Maranta F, Cianfanelli L, Cianflone D. Glycaemic Control and Vascular Complications in Diabetes Mellitus Type 2. Adv Exp Med Biol. 2021;1307:129–152.

[12] Crawford AL, Laiteerapong N. Type 2 Diabetes. Ann Intern Med. 2024;177(6): ITC81-ITC96.

[13] von Elm E, Altman DG, Egger M, Pocock SJ, Gotzsche PC, Vandenbroucke JP, et al. The Strengthening the Reporting of Observational Studies in Epidemiology (STROBE) statement: guidelines for reporting observational studies. Ann Intern Med, 2007, 147: 573–577.

[14] Zhou S, Chen R, Liu J, et al. Comparative Effectiveness and Safety of Atorvastatin Versus Rosuvastatin: A Multi-database Cohort Study. Ann Intern Med. 2024;177(12):1641–1651.

[15] Wu C, Zhang Y, Nie S, et al. Predicting in-hospital outcomes of patients with acute kidney injury. Nature communications. 2023, 14 (1): 3739.

[16] Xu X, Nie S, Xu H, et al. Detecting Neonatal AKI by Serum Cystatin C. J Am Soc Nephrol. 2023, 34 (7): 1253–1263.

[17] Alberti KG, Eckel RH, Grundy SM, et al. Harmonizing the metabolic syndrome: a joint interim statement of the International Diabetes Federation Task Force on Epidemiology and Prevention; National Heart, Lung, and Blood Institute; American Heart Association; World Heart Federation; International Atherosclerosis Society; and International Association for the Study of Obesity. Circulation. 2009; 20;120(16):1640-1645.

[18] Inker LA, Eneanya ND, Coresh J, et al. New Creatinine- and Cystatin C-Based Equations to Estimate GFR without Race. N Engl J Med. 2021, 385(19):1737–1749.

[19] Khunti K, Davies MJ. Clinical inertia-Time to reappraise the terminology? Prim Care Diabetes. 2017;11(1):105–106.

[20] Visseren FLJ, Mach F, Smulders YM, et al. 2021 ESC Guidelines on cardiovascular disease prevention in clinical practice. Eur Heart J. 2021;42(34):3227-3337.

[21] Austin PC, Lee DS, Fine JP. Introduction to the Analysis of Survival Data in the Presence of Competing Risks. Circulation. 2016;133(6):601–609.

[22] Bacchetti P. Current sample size conventions: flaws, harms, and alternatives. BMC Med. 2010;8:17.

[23] Zoungas S, Chalmers J, Neal B, et al. Follow-up of blood-pressure lowering and glucose control in type 2 diabetes. N Engl J Med. 2014;371(15):1392–1406.

[24] Cholesterol Treatment Trialists’ (CTT) Collaboration. The effects of lowering LDL cholesterol with statin therapy in people at low risk of vascular disease: meta-analysis of individual data from 27 randomised trials. Lancet. 2012;380 (9841):581-90.

[25] SPRINT Research Group. A Randomized Trial of Intensive versus Standard Blood-Pressure Control. N Engl J Med. 2015;373(22):2103–2116.

[26] Brunström M, Carlberg B. Association of Blood Pressure Lowering with Mortality and Cardiovascular Disease Across Blood Pressure Levels: A Systematic Review and Meta-analysis. JAMA Intern Med. 2018;178(1):28–36.

[27] Zoungas S, Patel A, Chalmers J, et al. Severe hypoglycemia and risks of vascular events and death. N Engl J Med. 2010;363(15):1410–1418.

[28] Gaba P, O’Donoghue ML, Park JG, et al. Association Between Achieved Low-Density Lipoprotein Cholesterol Levels and Long-Term Cardiovascular and Safety Outcomes: An Analysis of FOURIER-OLE. Circulation. 2023;147(16): 1192–1203.

[29] Kalantar-Zadeh K, Streja E, Kovesdy CP, et al. The obesity paradox and mortality associated with surrogates of body size and muscle mass in patients receiving hemodialysis. Mayo Clin Proc. 2010;85(11):991–1001.

[30] Kalantar-Zadeh K, Block G, Horwich T, et al. Reverse epidemiology of conventional cardiovascular risk factors in patients with chronic heart failure. J Am Coll Cardiol. 2004;43(8):1439–44.

[31] Ortega FB, Lavie CJ, Blair SN. Obesity and Cardiovascular Disease. Circ Res. 2016;118(11):1752–1770.

